# Bacterial Bloodstream Infections in Cameroon: A Systematic Review and Meta-Analysis of Prevalence and Antibiotic Resistance

**DOI:** 10.1101/2024.02.10.24302357

**Authors:** Moise Matakone, Patrice Landry Koudoum, Ravalona Jessica Zemtsa, Sen Claudine Henriette Ngomtcho, Isaac Dah, Michel Noubom

## Abstract

**Background:** The paucity of data on the epidemiology of bloodstream infection (BSI) in low and middle-income countries (LMICs) limits its effective prevention and management. This review sought to determine the prevalence, bacteriological and antimicrobial resistance profiles of bacteria implicated in BSI in Cameroon.

**Methods:** PubMed and Google Scholar databases were searched to identify relevant articles, which were screened according to the PRISMA guidelines. The data were analysed using comprehensive meta-analysis software. The I^2^ was used to evaluate heterogeneity between studies, Begg’s and Egger’s regression tests were used to evaluate publication bias, and random effects analysis was used to calculate the pooled prevalence.

**Results:** A total of 4223 blood cultures were obtained from the 10 included studies. The overall pooled prevalence of bacterial BSI was 26.31% (95% CI= 17.01%–38.35%). *Escherichia coli* (23.09%; 95% CI= 9.21%–47.05%), *Klebsiella* spp. (22.95%; 95% CI= 13.09%–37.07%), and *Staphylococcus aureus* (16.09%; 95% CI= 8.11%–29.43%) were the most common bacteria species. *E. coli* and *Klebsiella* spp. displayed the highest resistance to amoxicillin (82.65%; 95% CI= 63.25%–92.95% vs 86.42%; 95% CI= 55.90%–96.97%), amoxicillin + clavulanic acid (71.74%; 43.96–89.15% vs 73.06%; 95% CI= 38.70%–92.09%) and cotrimoxazole (76.22%; 95% CI= 51.33%–90.79% vs 65.81%; 95% CI= 45.08–81.86%). However, meropenem (26.73%; 95% CI= 20.76%–33.68%) and fosfomycin (14.85%; 95% CI= 9.07%–23.37%) were the least resistant in *E. coli* and *Klebsiella* spp., respectively. *Staphylococcus aureus* strains exhibited highest resistance to penicillin (84.37%; 95% CI= 68.13%–93.16%), erythromycin (44.80%; 95% CI= 33.37%–56.79%) and oxacillin (37.35%; 95% CI= 8.76%–78.74%) and lowest resistance to rifampicin (2.94%; 95% CI= 0.59%–13.39%), fusidic acid (6.73%; 95% CI= 2.55%–16.62%) and vancomycin (13.18%; 95% CI= 2.26%–49.86%).

**Conclusion:** This study reports a high prevalence of bacterial BSIs in Cameroon and the high resistance of these bacteria to common antibiotics. There is a pressing need to conduct BSI surveillance studies in all regions of Cameroon to generate data for evidence-based measures regarding BSI prevention and management.

**Prospero registration number:** CRD42023482760

## Background

Bloodstream infection (BSI) is characterized by positive blood cultures in patients with systemic symptoms and signs of infection [1]. BSI can eventually lead to sepsis, which is a complication characterised by life-threatening organ dysfunction caused by a dysregulated host response to infection [1]. Sepsis and BSI are significantly associated with morbidity and mortality worldwide and represent major public health concerns [2]. According to the World Health Organization (WHO), sepsis accounted for approximately 20% of all global deaths in 2017, affected 49 million people, and resulted in 11 million fatalities [2, 3]. Vulnerable populations such as pregnant women, elderly persons, patients with severe comorbidities, newborns, and those in low-resource settings are disproportionately affected [4, 5]. Cases of sepsis and sepsis-related deaths in low and middle-income countries (LMICs) account for 85% of the global incidence of sepsis [3] and hospital mortality due to sepsis is estimated to be 19% for moderate sepsis and 39% for severe sepsis in sub-Saharan Africa (SSA) [6]. Bacteria, especially *Enterobacterales*, which include *Escherichia coli*, *Klebsiella* spp., and *Salmonella enterica*, and gram-positive cocci, such as *Staphylococcus aureus* and coagulase-negative *Staphylococci*, are major species implicated in BSI and sepsis [7, 8].

Inappropriate use of antibiotics is known to exacerbate the burden of antimicrobial resistance (AMR) in bacterial pathogens [9]. Globally, there is an increase in resistance to front-line antibiotics, including β-lactams, fluoroquinolones, aminoglycosides, and macrolides [10, 11]. Therefore, AMR is a major public health concern and is expected to be the leading cause of mortality by 2050, with 10 million deaths per year if no immediate measures are taken [12]. AMR is a serious challenge in hospital settings because it worsens the effects of bacterial infections in general, especially sepsis, from extended hospital stays to increased mortality [13, 14].

In Cameroon, the bacterial BSI and sepsis and the AMR rate of implicated bacteria are still poorly documented. Although some studies have been published, summarizing these available data is important for providing information on the context-specific epidemiology of BSIs in terms of aetiology and AMR rate and for directing future BSI and sepsis prevention and management efforts in Cameroon. Thus, in this review, we estimated the pooled proportion of culture-confirmed bacterial BSIs and their antimicrobial resistance (AMR) rates in Cameroon.

## Methods

We conducted this systematic review and meta-analysis following the Preferred Reporting Items for Systematic Review and Meta-Analysis (PRISMA) 2020 guidelines [15], and the protocol was registered in the Prospero database (Ref: CRD42023482760, available from: https://www.crd.york.ac.uk/prospero/display_record.php?ID=CRD42023482760).

### Search strategy

The National Library of Medicine database (MEDLINE) was searched through PubMed, along with Google Scholar and hand search. These databases were searched to find all articles reporting the prevalence of culture-confirmed bacteria-driven bloodstream infections. Library research was performed using Medical Subject Headings (MeSH) and keywords paired with Boolean operators (**Figure S1 and Figure S2**).

**Figure 1:**
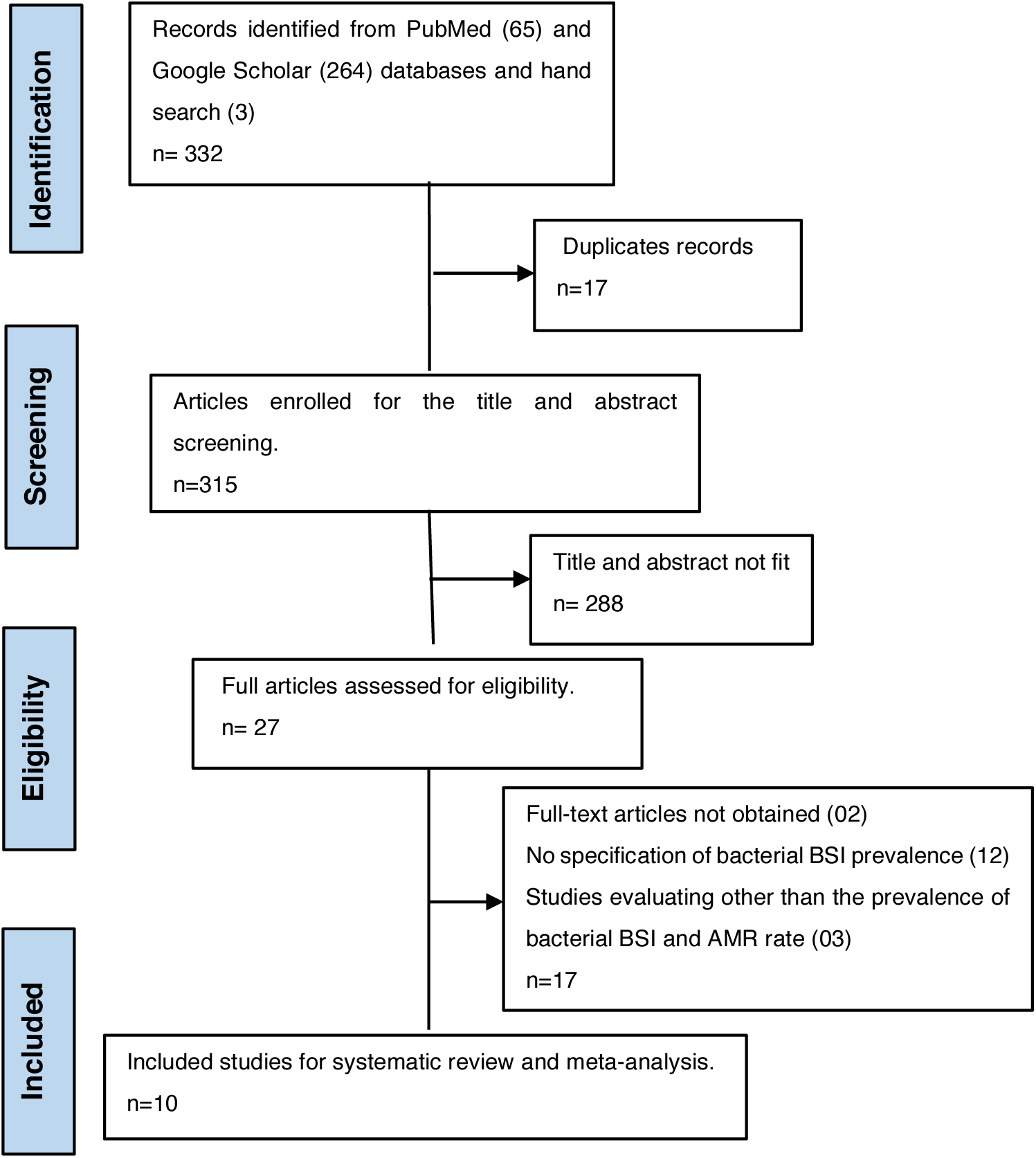
PRISMA flow chart of the study selection process

**Figure 2:**
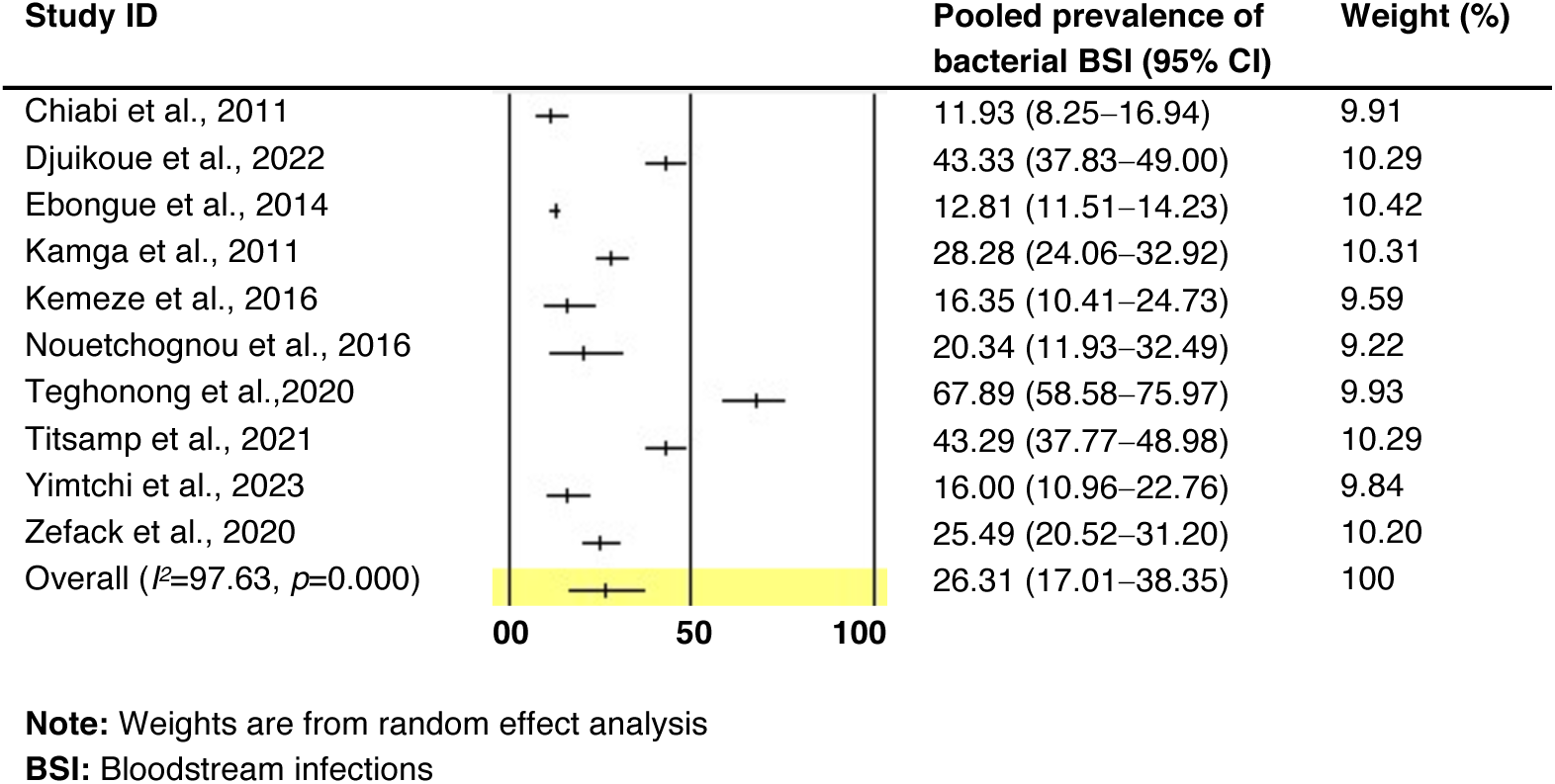
Forest plot of pooled prevalence of culture-confirmed bacterial bloodstream infections in Cameroon

### Eligibility criteria

Three authors independently searched the literature and evaluated the titles and abstracts of the papers. The articles included were studies that (1) reported culture-confirmed bacteria-driven bloodstream infections in Cameroon, (2) were published in peer-reviewed journals and (3) were published in English or French from 2010 to December 2023. Articles were excluded if (1) the full text was not assessable or (2) they were case reports or case series.

### Study selection

After the literature search, we uploaded all the articles to the EndNote (version 20.6) database, and duplicates were removed. Two reviewers screened the titles and abstracts to assess whether specific inclusion criteria were met and non-relevant and those that did not meet the inclusion criteria were excluded. Disagreements on whether an article should be included were resolved by a third reviewer.

### Data extraction

Two authors used a Microsoft Excel spreadsheet (version 2019) to extract the data from the included articles independently. The first author, publication year, region of study, study design, sample size, age of the study population, prevalence of BSIs, proportion of each bacteria causing BSIs and AMR rates of the bacteria groups were extracted. Inconsistencies were resolved through a group consensus.

### Quality assessment

The quality of the included studies was assessed using the Joanna Briggs Institute revised Critical Appraisal Checklist [16]. The checklist was applied to each study to assess the eight items, with each item awarded a point for “yes”, and the final score for each article was added and fell between zero and eight. Any disagreements among the authors were resolved by consensus. A study was considered low risk if it scored ≥ 4 points.

### Data analysis

We exported the extracted data to Comprehensive Meta-Analysis Software (version 3.0) for analysis. Begg and Egger’s regression tests were used to objectively assess publication bias, while a funnel plot was used to visualise this bias [17]. A p-value < 0.05 was considered to indicate statistical significance [18]. Cochrane Q statistics were used to assess the presence and magnitude of heterogeneity between studies. Heterogeneity was quantified using I^2^ expressed as a percentage. Values of 75, 50 and 25% were considered high, medium and low heterogeneity respectively [19].

## Results

### Results of the literature search

The online database search from PubMed and Google Scholar yielded 329 articles and three articles through the hand search. Duplicates were removed, and 315 articles were included in the title and abstract screening. Finally, 27 articles were screened for full-text review, and 10 that met the study objectives were included (**Figure 1**).

### Publication bias

When the BSI among study participants was analysed, a visual inspection of the funnel plot showed that there was no evidence of publication bias among the included studies (**Figure S3**). Similarly, the Egger’s regression intercept (4.89, 95% CI= −5.23–15.01, p= 0.298) and Begg test (p= 0.79) were not statistically significant.

### Characteristics of the included studies

A total of 4223 blood cultures were obtained from the 10 included studies, 920 (21.79%) of which were performed exclusively in the paediatric population (children under 2 years old). The studies were conducted in two of the 10 regions of Cameroon and included patients from only two cities, Douala (n=2547; 60.31%) [20–22] and Yaoundé (n=1676; 39.69%) [23–29]. Most of the studies were cross-sectional (n=08) [20, 22–25, 27–29], one was longitudinal [26] and one was retrospective [21], but all were of low risk. Only cross-sectional studies reported the age of the patients, which ranged from 0 to 92 years (**Table S1**).

**Table 1:** Pooled rate of major bacteria causing bloodstream infections in Cameroon.

| Bacterial species | Pooled rate (95%CI)<br>from random effect | I<sup>2</sup> | P-value | References |
| --- | --- | --- | --- | --- |
| *E. coli* | 23.09 (9.21–47.05) | 96.42 | 0.000 | [20, 21, 23-29] |
| *Klebsiella* spp. | 22.95 (13.09–37.07) | 83.02 | 0.000 | [20, 21, 24, 26-29] |
| *Enterobacter* spp. | 8.06 (4.96–12.83) | 31.74 | 0.186 | [20, 21, 24, 26-29] |
| *Salmonella enterica* | 7.15 (3.68–13.43) | 51.22 | 0.056 | [20, 21, 24, 26-29] |
| *Acinetobacter* spp. | 3.74 (2.07–6.66) | 23.80 | 0.247 | [20, 21, 24, 26-29] |
| *Pseudomonas* spp. | 3.17 (1.96–5.07) | 0.00 | 0.948 | [20, 21, 24, 26-29] |
| *Staphylococcus aureus* | 16.09 (8.11–29.43) | 81.05 | 0.000 | [20, 21, 24, 26-29] |
| *Coagulase negative Staphylococci* | 10.85 (3.15–31.28) | 93.82 | 0.000 | [20, 21, 24, 26-29] |
| *Streptococcus* spp. | 5.24 (2.68–10.00) | 50.33 | 0.060 | [20, 21, 24, 26-29] |

### Pooled prevalence and bacterial aetiology of bloodstream infections

The heterogeneity among the included studies was statistically significant (I^2^= 97.63%; p=0.000). Thus, we used a random effect model to calculate the pooled prevalence of culture-confirmed bacteria-driven bloodstream infections in Cameroon, and 95% confidence intervals (CIs) were represented on forest plots (**Figure 2**). Blood culture was performed for 4223 patients, and 888 were positive for a bacterial BSI. The bacterial BSI prevalence in this study ranged from 11.93% to 67.89%, and the overall pooled prevalence was 26.31% (95% CI= 17.01%–38.35%).

The main bacterial groups reported in this study were *Enterobacterales, Staphylococci* and *Streptococci* (**Table 1**). Among the *Enterobacterales* group, *Escherichia coli* (23.09%; 95% CI= 9.21%–47.05%), was the most common bacteria, followed by *Klebsiella* spp. (22.95%; 95% CI= 13.09%–37.07%), *Enterobacter* spp. (7.66%; 95% CI= 4.46%–12.83%) and *Salmonella enterica* (7.15%; 95% CI= 3.68%–13.43%). Seven studies reported gram-positive cocci, of which *Staphylococcus aureus* was the most common (16.09%; 95% CI= 8.11%–29.43%), followed by *coagulase-negative Staphylococci* (10.85%; 95% CI= 3.15%–31.28%) and *Streptococcus* spp. (5.24%; 95% CI= 2.68%–10.00%).

### Antimicrobial resistance rates of bacterial BSIs

This study reported only the pooled AMR rate of the most represented bacteria in each group. Overall, both *E. coli* and *Klebsiella* spp. displayed the highest resistance to amoxicillin (82.65%; 95% CI= 63.25%–92.95% vs 86.42%; 95% CI= 55.90%–96.97%), amoxicillin + clavulanic acid (71.74%; 43.96–89.15% vs 73.06%; 95% CI= 38.70%–92.09%) and cotrimoxazole (76.22%; 95% CI= 51.33%–90.79% vs 65.81%; 95% CI= 45.08–81.86%). Additionally, resistance to third and fourth-generation cephalosporins and monobactam was high among *E. coli* isolates: cefotaxime, 46.46% (95% CI= 26.77%–67.31%); ceftazidime, 49.92% (95% CI= 22.54%–70.32%); cefepime, 51.53% (95% CI= 40.92%–62.00%); aztreonam, 49.95% (95% CI= 40.59%–59.31%); *Klebsiella* spp isolates: cefotaxime, 40.98% (95% CI= 19.41%–66.69%); ceftazidime, 45.14% (95% CI= 16.76%–77.08%); and aztreonam, 54.25% (95% CI= 34.16%–73.04%). However, meropenem (26.73%; 95% CI= 20.76–33.66%) and fosfomycin (14.85%; 95% CI= 9.07%–23.37%) were the most effective against *E. coli* and *Klebsiella* spp., respectively (**Table 2**).

**Table 2:** Pooled antibiotic resistance rate of the most represented gram-negative bacilli isolated from bloodstream infections in Cameroon.

| Antibiotics | Pooled AMR rate from random model (95% CI) |  |  |  |  |  |
| --- | --- | --- | --- | --- | --- | --- |
|  | *E. coli* | I<sup>2</sup>(p-value) | N° isolates tested | *Klebsiella* spp. | I<sup>2</sup>(p-value) | N° studies |
| Amoxicillin | 82.65 (63.25–92.95) | 80.60 (0.000) | 275 | 86.42 (55.90–96.97) | 65.68 (0.002) | 107 |
| Amoxicillin + clavulanic acid | 71.74 (43.96–89.15) | 91.69 (0.000) | 276 | 73.06 (38.70–92.09) | 73.34 (0.024) | 99 |
| Cefotaxime | 46.46 (26.77–67.31) | 91.25 (0.000) | 263 | 40.98 (19.41–66.69) | 65.20 (0.056) | 99 |
| Ceftazidime | 49.92 (22.54–70.32) | 87.69 (0.000) | 274 | 45.14 (16.76–77.08) | 77.92 (0.011) | 99 |
| Cefepime | 51.53 (40.92–62.00) | 41.50 (0.181) | 187 | NA | NA | NA |
| Cefoxitin | 33.19 (18.55–52.01) | 82.56 (0.000) | 281 | 35.80 (19.48–56.25) | 46.74 (0.131) | 107 |
| Aztreonam | 49.95 (40.59–59.31) | 0.00 (0.806) | 108 | 54.25 (34.16–73.04) | 0.00 (0.521) | 24 |
| Meropenem | 26.73 (20.76–33.68) | 0.00 (0.501) | 180 | NA | NA | NA |
| Amikacin | 37.54 (18.66–61.15) | 89.07 (0.000) | 274 | 26.75 (6.12–67.16) | 76.10 (0.009) | 107 |
| Gentamycin | 49.48 (37.34–61.68) | 61.89 (0.022) | 281 | 50.07 (31.65–68.48) | 47.64 (0.106) | 115 |
| Ciprofloxacin | 41.73 (32.62–51.45) | 21.87 (0.275) | 191 | 56.52 (29.94–79.81) | 62.49 (0.031) | 115 |
| Fosfomycin | NA | NA | NA | 14.85 (9.07–23.37) | 0.00 (0.728) | 99 |
| Nalidixic acid | 34.60 (28.10–41.73) | 0.00 (0.968) | 185 | NA | NA | NA |
| Cotrimoxazole | 76.22 (51.33–90.79) | 84.67 (0.000) | 275 | 65.81 (45.08–81.86) | 56.12 (0.077) | 107 |

*Staphylococcus aureus* was highly resistant to penicillin (84.37%; 95% CI= 68.13%–93.16%), erythromycin (44.80%; 95% CI= 33.37%–56.79%) and oxacillin (37.35%; 95% CI= 8.76%–78.74%). However, a low percentage of *Staphylococcus aureus*-resistant strains was detected for non-β-lactam antibiotics, including rifampicin (2.94%; 95% CI= 0.59%–13.39%), fusidic acid (6.73%; 95% CI= 2.55%–16.62%), and vancomycin (13.18%; 95% CI= 2.26%–49.86%). In the *CONS* group, a high percentage of strains were resistant to penicillin (89.91%; 95% CI= 65.95%–97.62%), cotrimoxazole (56.88%; 95% CI= 31.93%–78.78%) and oxacillin (50.70%; 95% CI= 24.97%–76.07%), while rifampicin (11.13%; 95% CI= 4.46%–25.16%) and cefoxitin (14.87%; 95% CI= 1.08%–73.62%) (**Table 3**).

**Table 3:** Pooled antibiotic resistance rate of the most represented Staphylococci causing bloodstream infections in Cameroon.

| Antibiotics | Pooled AMR rate from random model (95% CI) |  |  |  |  |  |
| --- | --- | --- | --- | --- | --- | --- |
|  | *Staphylococcus aureus* | I<sup>2</sup>(p-value) | N° isolates tested | *Coagulase negative Staphylococci* | I<sup>2</sup>(p-value) | N° studies |
| Penicillin | 84.37 (68.13–93.16) | 39.86 (0.197) | 61 | 89.91 (65.95–97.62) | 66.56 (0.050) | 93 |
| Oxacillin | 37.35 (8.76–78.74) | 88.19 (0.004) | 61 | 50.70 (24.97–76.07) | 83.81 (0.002) | 93 |
| Cefoxitin | 18.85 (1.16–82.07) | 81.51 (0.020) | 28 | 14.87 (1.08–73.62) | 85.97 (0.008) | 71 |
| Gentamycin | 18.85 (1.16–8.3) | 81.51 (0.020) | 28 | 36.67 (18.66–59.37) | 76.23 (0.015) | 93 |
| Erythromycin | 44.80 (33.37–56.79) | 0.00 (0.892) | 67 | 46.81 (27.28–67.37) | 73.82 (0.022) | 93 |
| Lincomycin | 25.69 (3.96–74.33) | 90.21 (0.001) | 61 | 42.05 (32.24–52.53) | 2.26 (0.359) | 93 |
| Rifampicin | 2.94 (0.59–13.39) | 0.00 (0.450) | 61 | 11.13 (4.46–25.16) | 40.10 (0.196) | 51 |
| Tetracycline | 28.04 (5.81–71.12) | 84.06 (0.002) | 67 | 41.71 (26.95–58.12) | 56.10 (0.103) | 93 |
| Cotrimoxazole | 27.26 (14.90–44.51) | 35.99 (0.211) | 61 | 56.88 (31.93–78.78) | 81.37 (0.005) | 93 |
| Vancomycin | 13.18 (2.26–49.86) | 66.72(0.050) | 67 | 30.40 (14.37–53.20) | 63.64 (0.064) | 93 |
| Fusidic acid | 6.73 (2.55–16.62) | 0.00(0.637) | 61 | 25.46 (17.44–35.58) | 1.29 (0.363) | 93 |

## Discussion

This review aimed to determine the pooled prevalence of bacterial BSIs and the AMR rates of implicated bacteria in Cameroon. The 10 studies selected for this review described 4223 blood cultures, and the overall pooled prevalence of BSI was 26.31%. This prevalence is higher than those reported in Africa by Reddy *et al*. (2010) and in Africa and Asia by Marchello *et al.* (2019), which were 7.4% and 10.8%, respectively [30, 31]. However, studies conducted in Ethiopia by Bitew *et al.* (2023), Legese *et al.* (2022) and Abebe *et al.* (2021) documented a similar prevalence of BSI [32–34]. The observed difference in this review may be due to the small sample size of many included studies [20, 22, 26, 27] and their study design.

The study revealed that *Enterobacterales*, including *Escherichia coli* (23.09%) and *Klebsiella* spp. (22.95%), were the leading bacteria implicated in BSI. Studies conducted in several low and middle-income countries have demonstrated the strong implication of these bacteria in bloodstream infections and sepsis, especially in hospital settings [10, 34, 35]. However, these results contradict those recorded by Wattal *et al.* (2020) and Reddy *et al.* (2010), where *Salmonella* spp. were the leading cause of BSI and sepsis [11, 30]. This contradiction might be due to the differences in the bacterial identification methods used, which are poorly described in some studies and can vary considerably depending on the laboratory [20, 21, 26]. Moreover, the observed contrast may be due to the differences in origins (community vs hospital) of bacteria driving BSIs. *Salmonella* spp. has been highly associated with community-acquired BSIs while *Escherichia coli* and *Klebsiella* spp. are to hospital-acquired BSIs [7, 30]. This may suggest that a greater share of BSIs reported in this review is of nosocomial origin.

The most frequent *Enterobacterales* reported in this review exhibited high resistance to β-lactam antibiotics, especially third- and fourth-generation cephalosporins and monobactam. This might indicate the production of β-lactamases, particularly extended-spectrum β-lactamase enzymes, by most of these bacteria even though most of the included studies failed to report resistance phenotypes.

## Limitations

The statements and conclusions in this review cannot be generalised to Cameroon, as the included studies investigated only two regions and some were limited by small sample sizes. Additionally, the study was unable to determine whether the BSI was community or hospital-acquired. Finally, the study failed to report AMR among several bacteria or the resistance phenotypes of the implicated bacteria. Notwithstanding, this review provides the most comprehensive data regarding bacterial BSI prevalence and AMR epidemiology in Cameroon.

## Conclusion

The prevalence of bacterial bloodstream infection in this study was elevated. *Enterobacterales*, including *Klebsiella* spp. and *Escherichia coli*, and gram-positive cocci, particularly *Staphylococcus aureus*, remain the leading causes of bloodstream infection. Antibiotic resistance was greater for β-lactams in both *Enterobacterales* and gram-positive cocci. This study may help clinicians optimise their antibiotic prescription protocols for the early management of bacterial bloodstream infections in the absence of antibiotic susceptibility testing results. This review also calls for researchers to expend more effort to produce AMR data to permit policymakers and stakeholders to better manage and prevent bloodstream infection in Cameroon.

## Data Availability

All data produced in the present work are contained in the manuscript

## List of abbreviations

AMR: Antimicrobial Resistance
BSI: Bloodstream Infections

## Declarations

### Ethics approval and consent to participate

Not applicable

### Consent for publication

Not applicable

### Availability of data and materials

The datasets generated and/or analysed during the current study are available from the corresponding author upon reasonable request.

### Author contributions

MM conceived and designed the study. MM and PLK coordinated the literature search. PKL, RJZ and MM undertook the data extraction, quality assessment and statistical analysis. MM, PLK, and RJZ prepared the first draft of the manuscript. ID, SCHN and MN critically reviewed the manuscript. All the authors read and approved the final version of the manuscript.

## Acknowledgements

Not applicable

## Supplementary figures

**Figure S1:**
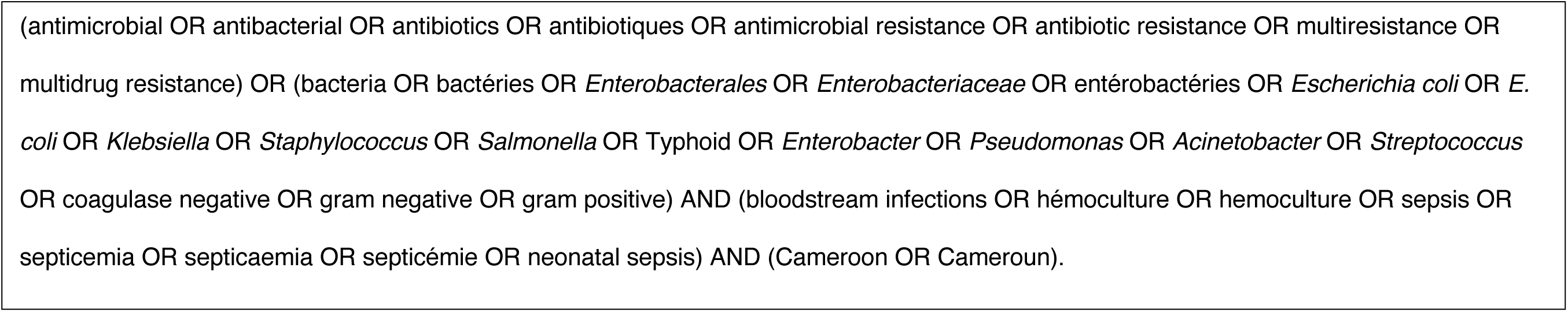
Search terms used to identify relevant literature from PubMed

**Figure S2:**
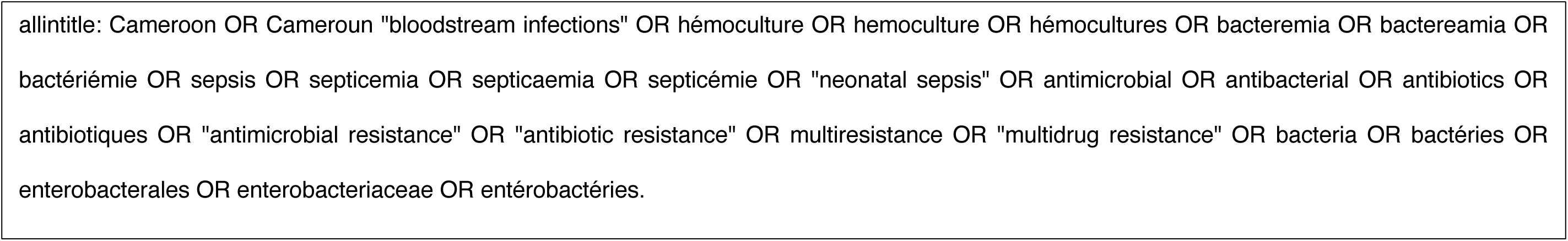
Search terms used to identify relevant literature from Google Scholar

**Figure S3:**
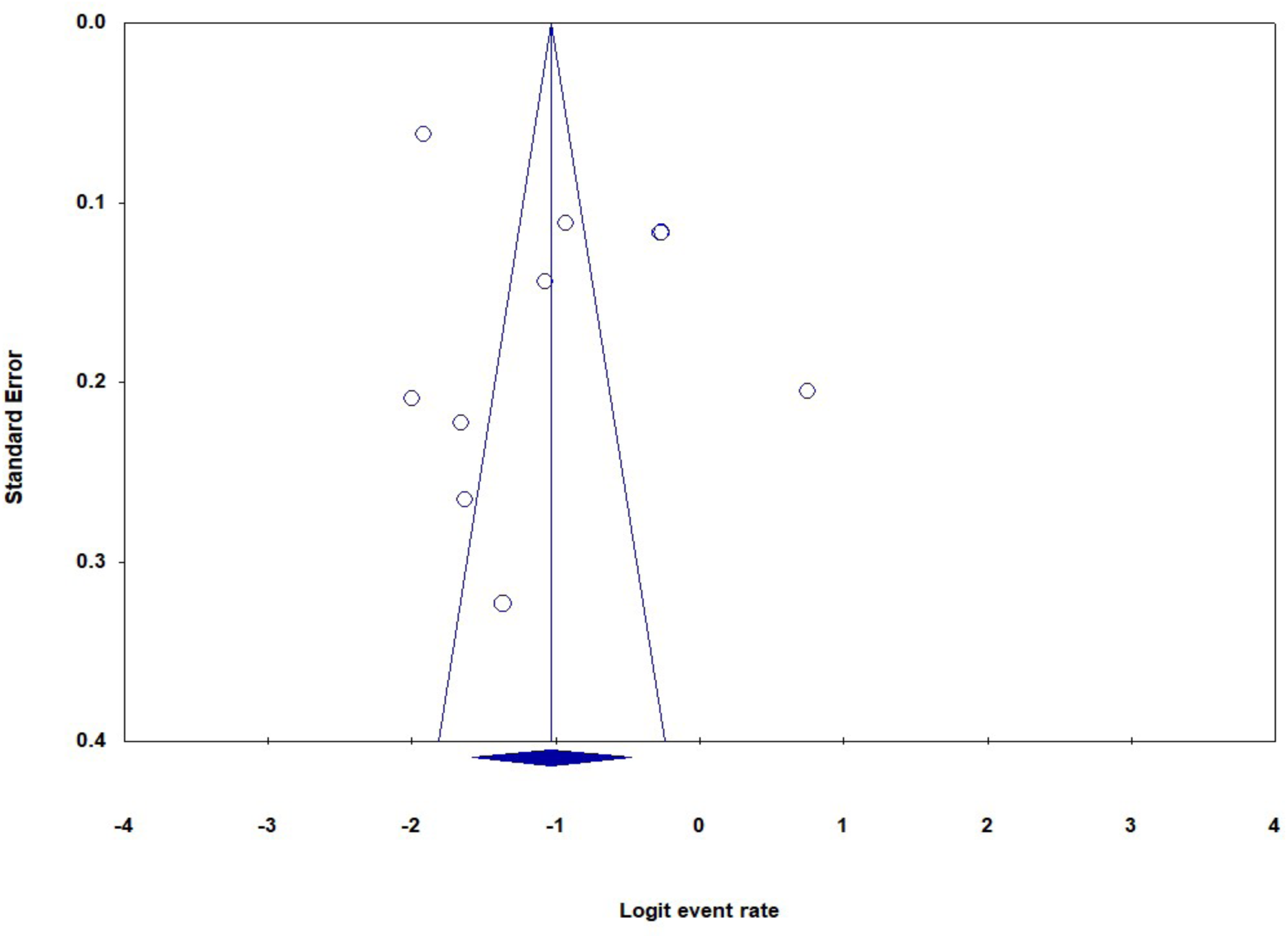
Funnel plot of standard error by logit event rate

**Figure S4:**
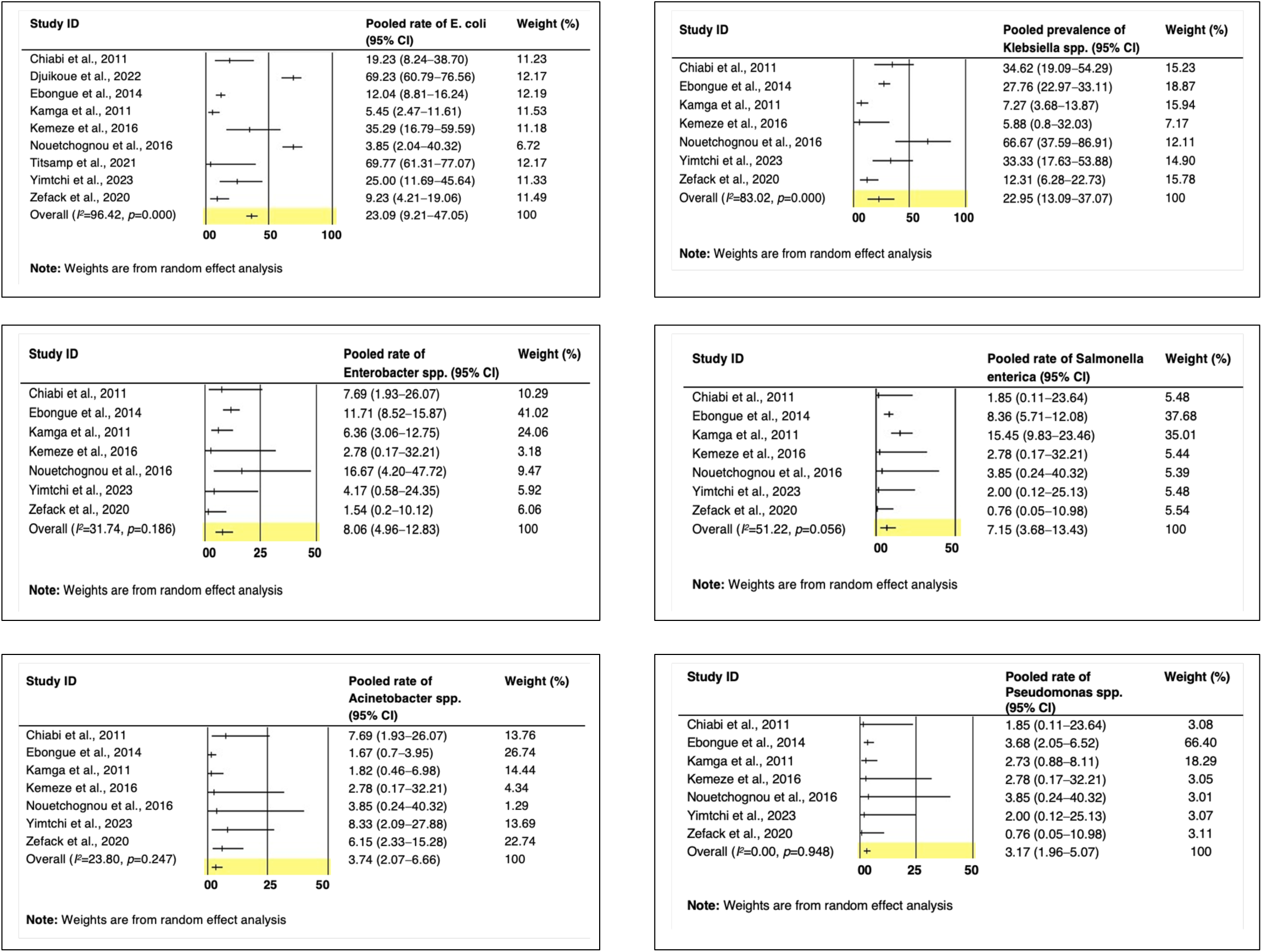
Forest plots of pooled rates of major gram-negative bacilli causing bloodstream infections in Cameroon

**Figure S5:**
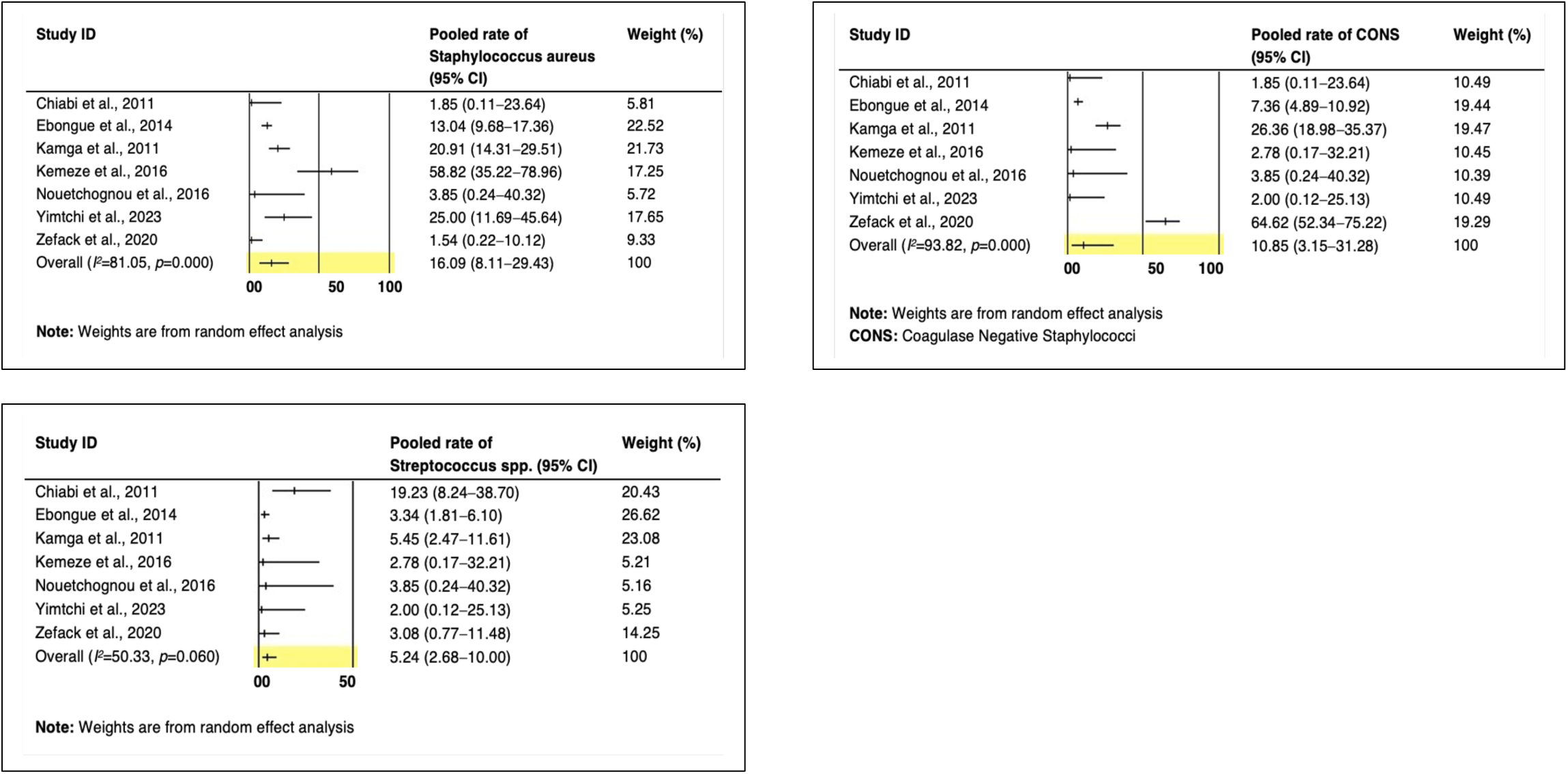
Forest plots of pooled rates of major gram-positive cocci causing bloodstream infections in Cameroon

## Supplementary table

**Table S1:**
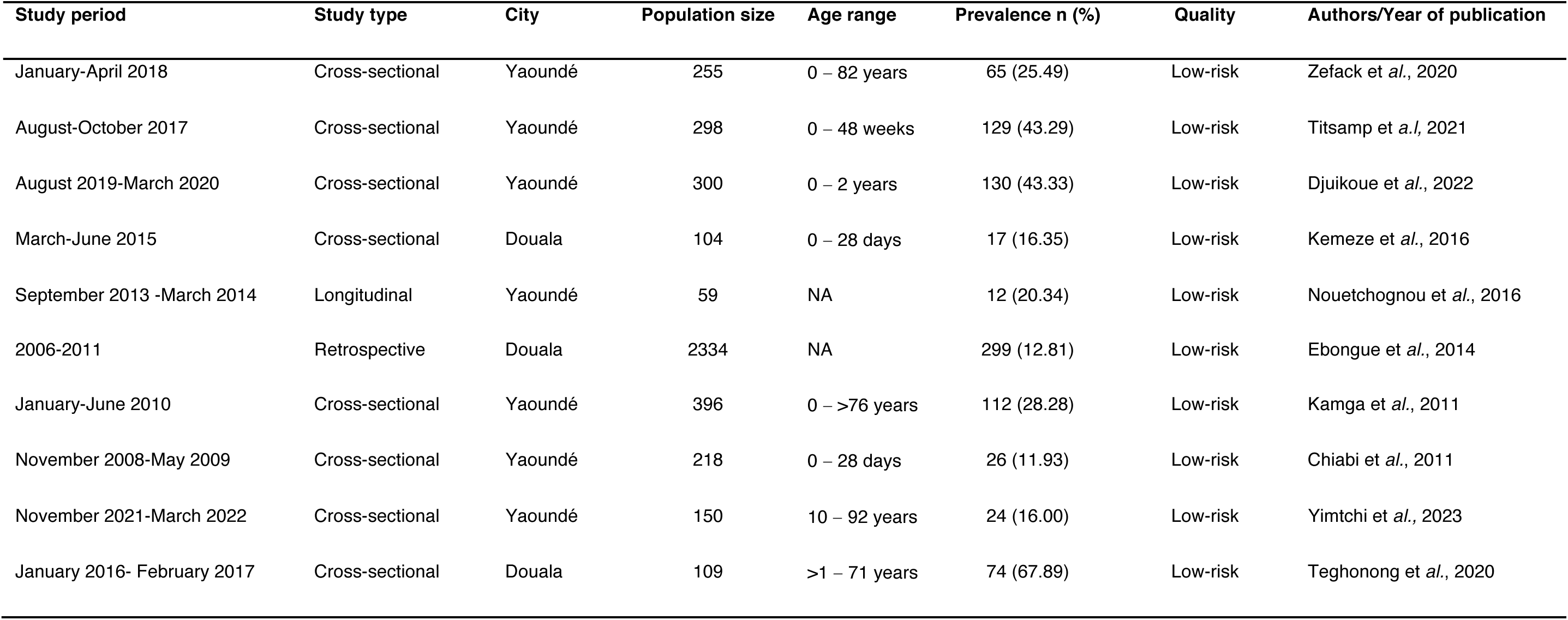
Summary characteristics of studies included in the meta-analysis.

